# Randomized controlled trial of convalescent plasma therapy against standard therapy in patients with severe COVID-19 disease

**DOI:** 10.1101/2020.11.02.20224303

**Authors:** Manaf AlQahtani, Abdulkarim Abdulrahman, Abdulrahman Almadani, Salman Yousif Alali, Alaa Mahmood Al Zamrooni, Amal Hamza Hejab, Ronán M Conroy, Pearl Wasif, Stephen L Atkin, Sameer Otoom, Manal Abduljalil

**Author notes:** Corresponding author: Manaf AlQahtani, Bahrain Defence Force Hospital, Bahrain. Clinical trials registration. NCT04356534.

## Abstract

**Background:** Convalescent plasma (CP) therapy in COVID-19 disease has been suggested to improve clinical outcome in severe disease. This pilot study was designed to inform the design of a definitive phase 3 clinical trial.

**Methods:** This was a prospective, interventional and randomized open label pilot trial involving 40 patients with COVID-19 who were requiring oxygen therapy and who had radiological evidence of pneumonia. Twenty COVID-19 patients received two 200ml transfusions of convalescent patient CP over 24 hours were compared with 20 patients who received routine care alone. The primary outcome was the requirement for ventilation. The secondary outcomes were white blood cell count, lactate dehydrogenase (LDH), C-reactive protein (CRP), Troponin, Ferritin, D-Dimer, procalcitonin, mortality rate at 28 days.

**Results:** The CP group were a higher risk group with higher ferritin levels (p<0.05) though respiratory indices did not differ. The primary outcome measure – ventilation – was required in 6 controls and 4 patients on CP (risk ratio 0.67 95% CI 0.22 – 2.0, p=0.72); mean time on ventilation was 10.5 days in the control against 8.2 days in patients on CP (p=0.81). There were no differences in secondary measures at the end of the study. Two patients died in the control and one patient in the CP arm.

**Conclusion:** There were no significant differences in the primary or secondary outcome measures between CP and standard therapy though fewer patients required ventilation and for a shorter period of time. The study showed that CP therapy appears to be safe and it is feasible to perform a definitive phase 3 clinical trial using this study protocol.

## Introduction

Coronavirus disease 2019 (COVID-19) is caused by the severe acute respiratory syndrome coronavirus 2 (SARS-CoV-2) and has developed into a pandemic with serious global public health and economic sequelae. As of October 31st 2020, more than 46 million cases have been confirmed worldwide leading to over 1,100,000 deaths^1^. There is no vaccine available, though several are currently in development in clinical trials. There have been a number of reports of medication such as remdesivir that have shown efficacy with shorter time to recovery having antiviral properties with efficacy against SARS-CoV-2. ^2^

Plasma therapy using Convalescent Plasma (CP) transfusion refers to a form of passive immunization, where neutralizing antibodies from a recovered donor are injected in the infected patient with the aim of altering the course of the disease. This has been shown to be effective in severe acute respiratory syndrome^3^, Ebola virus infection ^4^ and in H1N1 influenza^5^. More recently there has been a report of the use of CP in the treatment of 5 ventilated COVID-19 patients with the suggestion of expedited recovery as the patients improved 1 week after the transfusion^6^. A randomized trial with CP therapy in severely and critically ill COVID-19 patients undertaken in China was stopped early as recruitment slowed own due to the decrease in COVID-19 cases in China^7^. An open label Expanded Access Program in the US studied the effect on CP on mortality in more than 35,000 participants with COVID-19 that concluded that CP transfusion with high antibody level and early during admission was associated with decreased mortality^8^. However, no studies have been undertaken with CP therapy in hypoxic patients and therefore this pilot trial was undertaken to inform the design of a definitive study.

## Methods

This was a prospective, randomized, controlled open label pilot study involving 40 patients with severe COVID-19 disease confirmed by RT-PCR testing^9^. All patients gave written informed consent. This study was approved by the National COVID-19 Research Committee, and the Bahrain Defence Force Hospital Ethics committee and was conducted in accordance with the Declaration of Helsinki and local regulations. The trial was registered in clinicaltrials.gov with registration number NCT04356534. The trial was conducted upon the approved protocol by the National Research and Ethics committee. The study protocol is attached in the Appendix.

### Participants

Patients were recruited from two medical centres. The study recruitment was from April, 2020, to June, 2020.

### Inclusion Criteria

Inclusion criteria were: (1) signed informed consent; (2) aged at least 21 years; (3) COVID-19 diagnosis based on polymerase chain reaction (PCR) testing; (5) Hypoxia (Oxygen saturation of less than or equal 92% on air, or PO2 < 60mmHg in arterial blood gas, or arterial partial pressure of oxygen (PaO)/fraction of inspired oxygen (FIO) of 300 or less) and patient requiring oxygen therapy (6) pneumonia confirmed by chest imaging.

### Exclusion Criteria

Exclusion criteria were the following: (1) Patients with mild disease not requiring oxygen therapy; (2) Patients with normal CXR or CT scan; (3) Patients requiring ventilatory support (invasive or non-invasive); (4) Patients with a history of allergy to plasma, sodium citrate or methylene blue, or those with a history of autoimmune disease or selective IGA deficiency.

### Randomization

Following informed consent and screening, the patients were block randomised (in blocks of 4) by computer-generated random numbering to either the standard therapy or CP arms (Figure 1).

**Figure 1.**
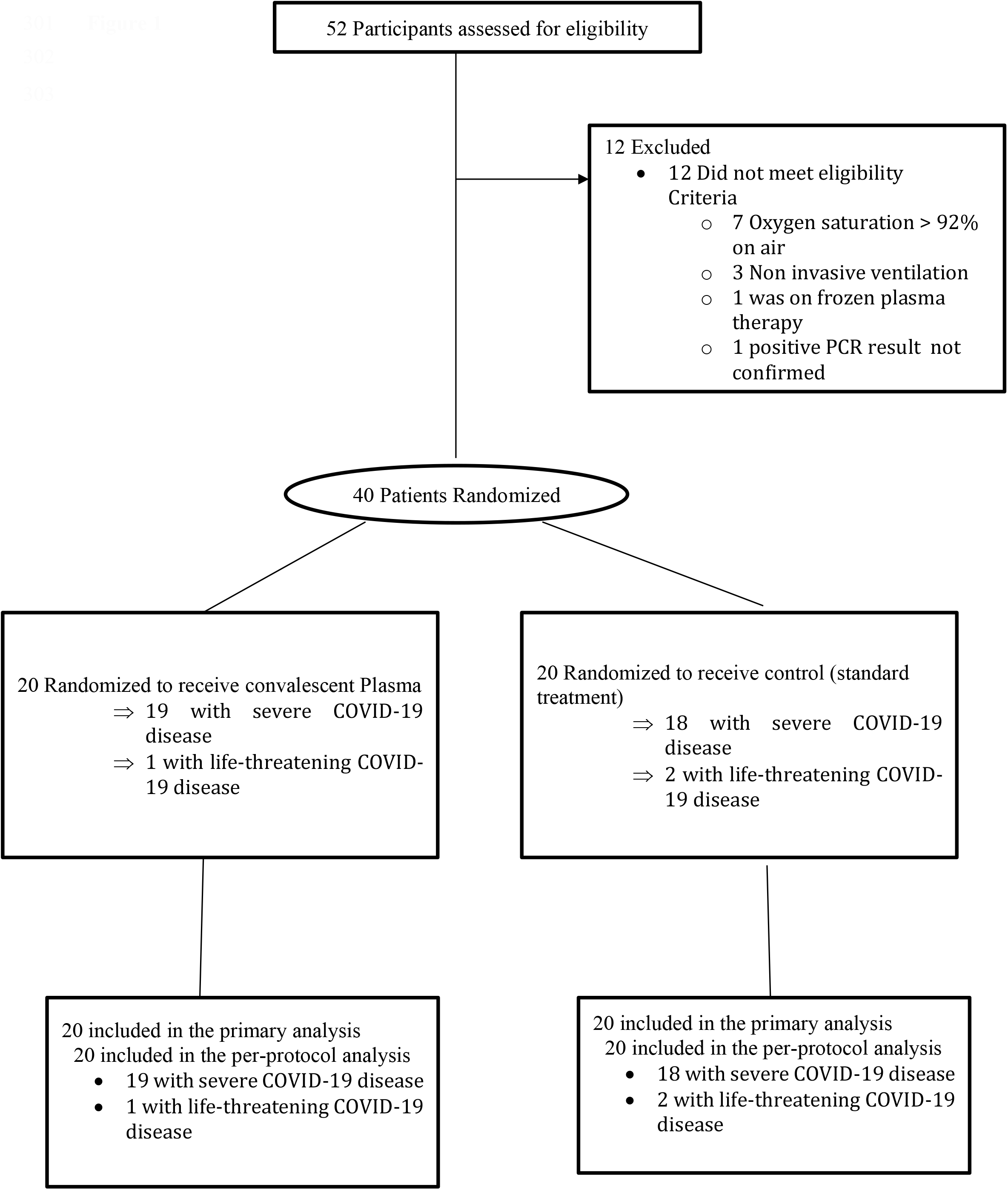

Patients and clinicians were not blinded to the treatment given.

### Convalescent Plasma donors

Patients who had recovered from COVID-19 and had been discharged from hospital for more than 2 weeks were approached to be volunteer donors. The criteria for donors included (1) ability to give informed consent; (2) men or nulliparous women (all women had a pregnancy test except for postmenopausal women); (3) PCR COVID-19 negative from respiratory tract; (4) patients were symptom free; (5) patients above the ages of 21; (6) body weight more than 50kg; (7) met all donor selection criteria employed for routine plasma collection and plasmapheresis procedures at the collection centre. Convalescent plasma collection was performed followed by plasma extraction as detailed.

COVID-19 CP collection and handling was undertaken at Bahrain Defence Force Hospital Blood Bank. History and demographics were obtained from each of the plasma donors who met all donor selection criteria employed for routine plasma collection and plasmapheresis procedures at the collection center. Approximately 450 ml of whole blood was venesected together with testing for anti-SARS-CoV-2 antibody titres.

Whole blood units were separated into packed red blood cells and plasma; the plasma was removed and stored at ≤ −18 °C. Laboratory tests including blood group (ABO/Rh), antibody screening, blood phenotype and transfusion-transmissible infections screening (which includes hepatitis B virus, hepatitis C virus, HIV, and syphilis). Convalescent compatible plasma was thawed and stored at 2°C - 6°C for 24 hours. Antibody levels were measured using Lansionbio COVID-19 IgM/IgG Test kit (Lansion Biotechnology Co., Ltd, China).

### Convalescent Plasma Transfusion

ABO matched CP units were selected for transfusion and transfused into the COVID-19 patients using standard clinical transfusion procedures.

The dosage of CP was be 400 mL, given as 200ml over 2hrs over 2 successive days; the infusion rate was monitored and amended if there was a risk of fluid overload.

Patients prior to CP therapy were on standard supportive treatment including control of fever (paracetamol) and possible therapy including antiviral medications, Tocilizumab and antibacterial medication.

### Standard Supportive Treatment

The standard supportive treatment included control of fever (paracetamol) and possible therapy including antiviral medications, Tocilizumab and antibacterial medication.

### Primary Outcome Measure

The primary end points were requirement for invasive or non-invasive ventilation, and, in patients who required ventilation, the duration of ventilation.

### Secondary Outcome Measures

Secondary outcome measures included C reactive protein, procalcitonin, lactate dehydrogenase, troponin, ferritin, D-Dimer, brain natriuretic peptide, lactate changes and 28day mortality rate

### Laboratory measurements

White blood cell count was measured by flow cytometry, lactate dehydrogenase (LDH) was measured using a kinetic method, C-reactive protein (CRP) and D-Dimer measured by an immuno-turbidimetric assay, Troponin, Ferritin and procalcitonin were measured by an electrochemiluminescence immunoassay according to the manufacturers’ instructions.

### Secondary analysis

Measurement of the effect of early CP transfusion (less than 3 days from admission) compared to late transfusion for the requirement of ventilation was undertaken. Further analysis was done to compare the mean antibody level of the CP on the requirement for ventilation.

### Statistical analysis

Power and sample size for pilot studies was based on Birkett and Day^10^. They concluded that a minimum of 20 degrees-of-freedom was required to estimate effect size and variability for normally distributed variables and hence 20 patients per group were recruited in this study; no allowance was made for dropouts, who are unlikely to be an issue in a study of this sort.

Baseline continuously distributed data are presented as means and standard deviations unless the data are skewed in which case the data median (25th/75th centiles) are presented; categorical data are shown by number and percent. Intention to treat analysis was used throughout.

For all statistical analyses, a two-tailed P <0.05 is considered to indicate statistical significance. Time to event analysis and logistic regression are used to analyse event-related outcomes. Effect size for secondary outcomes is shown as the Hodges-Lehmann median difference and its associated confidence interval, calculated using the community-contributed Stata command cendiff^11^. It should be noted that this is the median of all pairwise differences between the groups and does not correspond to the difference between the medians of the two groups.

Statistical analysis were performed using the Stata (StataCorp. College Station, TX: StataCorp LP, USA).

## Results

The participant demographic, clinical and biochemical characteristics are shown in Table 1 and medication after randomization is shown in Table 2. The two groups showed similar baseline epidemiological characteristics. The CP group showed higher ferritin levels (p=0.049); however, respiratory indices did not differ. There were more men than women, reflecting the demographics of the workforce in Bahrain.

**Table 1.** Subject demographics, clinical and laboratory characteristics at baseline.

| Factor | Level | Control | Plasma Arm | p-value |
| --- | --- | --- | --- | --- |
| N |  | 20 | 20 |  |
| Age, mean (SD) |  | 50.7 (12.5) | 52.6 (14.9) | 0.66 |
| Sex | Men | 15 (75%) | 17 (85%) | 0.43 |
|  | Women | 5 (25%) | 3 (15%) |  |
| Smoker |  | 0 (0%) | 0 (0%) |  |
| Diabetes |  | 9 (45%) | 7 (35%) | 0.52 |
| Hypertension |  | 5 (25%) | 5 (25%) | 1.0 |
| Cardiac diseases |  | 2 (10%) | 2 (10%) | 1.0 |
| Chronic kidney disease |  | 1 (5%) | 1 (5%) | 1.0 |
| Chronic lung disease |  | 0 (0%) | 3 (15%) | 0.072 |
| Chronic Liver disease |  | 0 (0%) | 0 (0%) |  |
| Oxygenation device required on admission | Nasal Cannula or face mask | 19 (95%) | 17 (85%) | 1.0 |
|  | Nonrebreather mask or High Flow Nasal Cannula | 1 (5%) | 3 (15%) |  |
| PaO<sub>2</sub>:Fio<sub>2</sub>, mean (SD) |  | 232 (56.8) | 220 (60.9) | 0.52 |
| Labs on admission |  |  |  |  |
| WBC, mean (SD) |  | 7.0 (4.0) | 5.9 (2.0) | 0.27 |
| LDH (N=35), mean (SD) |  | 345 (91.1) | 420 (172.2) | 0.11 |
| CRP, mean (SD) |  | 91 (52) | 110 (63) | 0.31 |
| D-Dimer (N=25), mean (SD) |  | 0.5 (0.2) | 1.3 (1.3) | <0.001** |
| Ferritin (N=39), mean (SD) |  | 631 (460) | 1045 (935) | 0.049** |
| Steroids | Yes | 4 (20%) | 1 (5%) | 0.15 |
| ** Wilcoxon Mann-Whitney test |  |  |  |  |

**Table 2.** Medication given after randomization.

| **Medication** | **Control** | **Plasma** |
| --- | --- | --- |
| Hydroxychloroquine | 20 | 17 |
| Lopinavir/ritonavir | 17 | 17 |
| Ribavirin | 7 | 5 |
| Azithromycin | 18 | 17 |
| Peginterfeon | 5 | 7 |
| tocilizumab | 6 | 6 |
| Methyl Prednisolone | 4 | 1 |
| Antibiotics | 20 | 20 |
| Anticoagultaion (LMWH/Heparin) | 20 | 19 |
| PPI | 12 | 7 |
| ACEi/ARB | 4 | 3 |
| Calcium Channel blocker | 3 | 5 |
| Beta blocker | 3 | 4 |
| Aspirin | 6 | 3 |
| Diuretics | 6 | 4 |
| Statin | 3 | 0 |
| Insulin | 6 | 8 |
| Metformin | 5 | 2 |
| Other Oral antidiabetic | 2 | 0 |
| Carbimazole | 1 | 0 |
| Thyroxine | 1 | 1 |
| Allopurinol | 1 | 0 |
| Acetylcysteine | 2 | 1 |

A total of 6 controls (30%) and 4 CP patients (20%) were ventilated (risk ratio 0.67 95%CI 0.22 – 2.0, p=0.72). Primary outcome measure, time to ventilation was not different (P=0·52, logrank test; Figure 2); time on ventilation did not differ (10.5 days control; 8.25 days CP (exact p=0.809)). Steroids were used in 3 control patients and none of the CP patients with no difference between groups (P=0·342). To detect a difference at 90% power, alpha 0.05, with a risk ratio of 0.7 and risk in control 20%, a sample size of 822 in each group would be needed.

**Figure 2.**
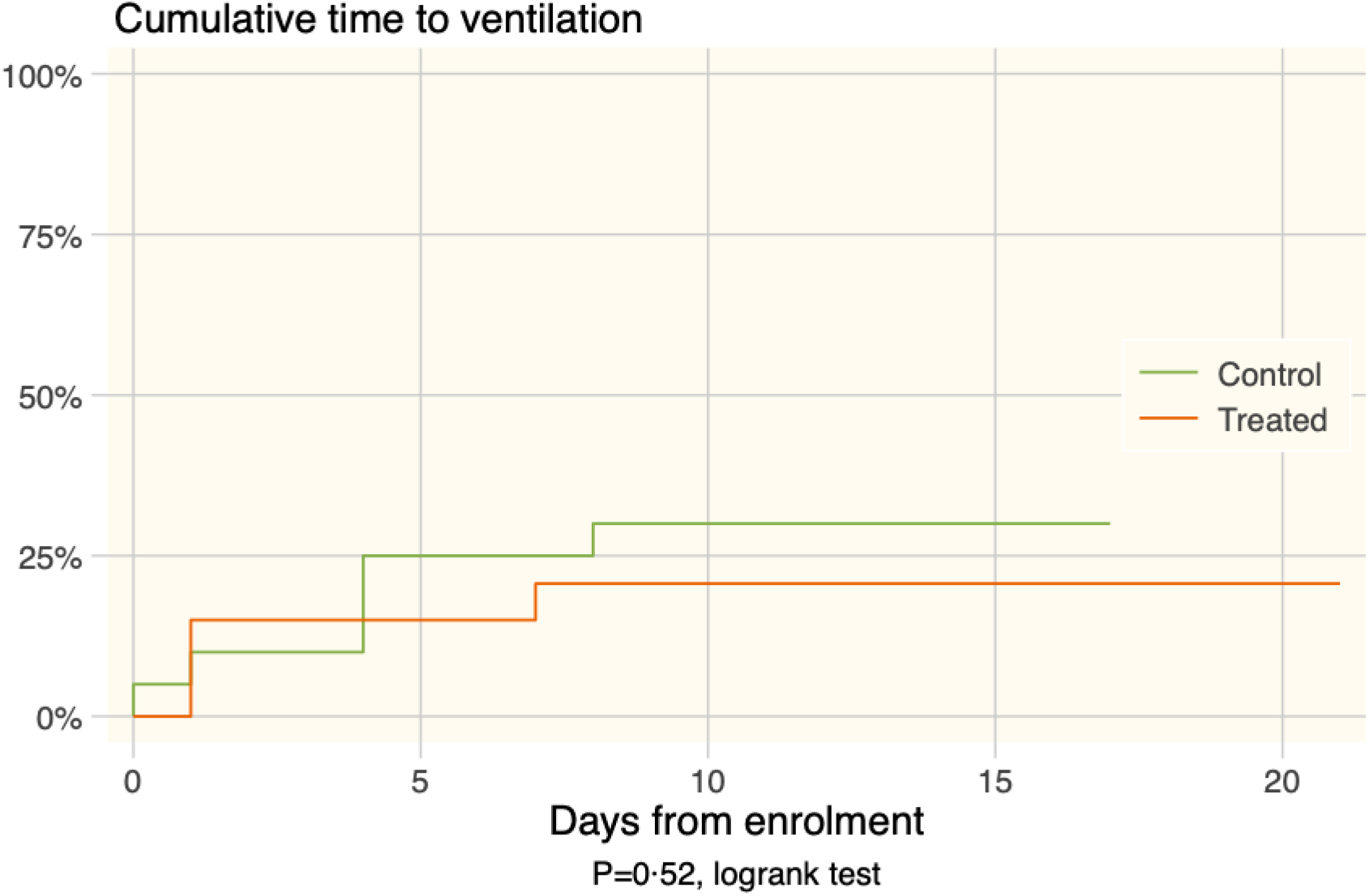
The plot shows cumulative ventilation rates for each group:

Table 3 shows the medians of secondary outcomes at discharge in patients who were discharged alive (18 control and 19 CP). The table also shows the Hodges Lehman median differences between the groups, with their associated confidence intervals. Significance levels are based on the Wilcoxon Mann-Whitney rank sum test.

**Table 3.**
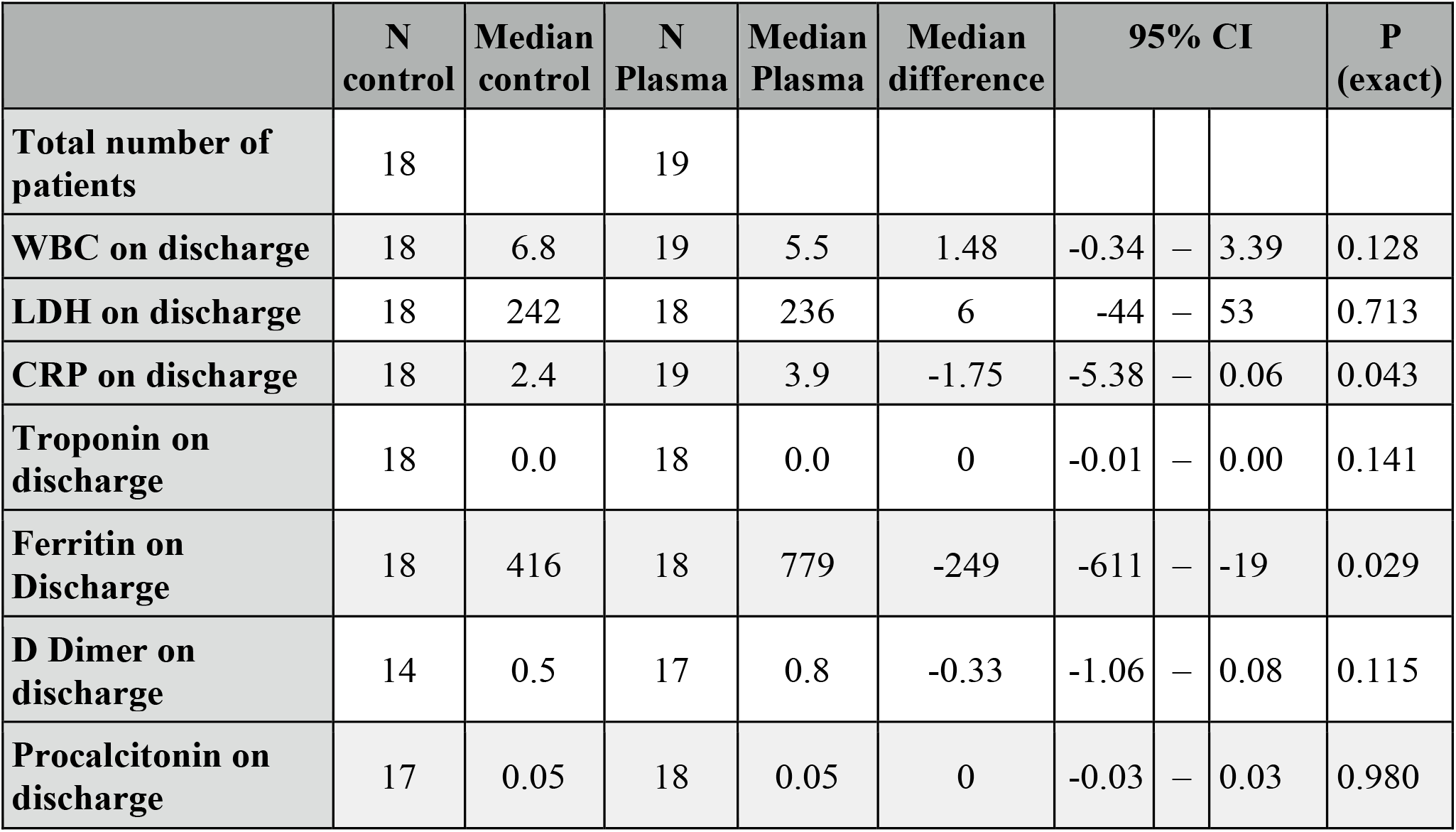
Secondary outcome measures between plasma (n=20) and control patients (n=20)

|  | N<br>control | Median<br>control | N<br>Plasma | Median<br>Plasma | Median<br>difference | 95% CI |  |  | P<br>(exact) |
| --- | --- | --- | --- | --- | --- | --- | --- | --- | --- |
| Total number of patients | 18 |  | 19 |  |  |  |  |  |  |
| WBC on discharge | 18 | 6.8 | 19 | 5.5 | 1.48 | -0.34 | – | 3.39 | 0.128 |
| LDH on discharge | 18 | 242 | 18 | 236 | 6 | -44 | – | 53 | 0.713 |
| CRP on discharge | 18 | 2.4 | 19 | 3.9 | -1.75 | -5.38 | – | 0.06 | 0.043 |
| Troponin on discharge | 18 | 0.0 | 18 | 0.0 | 0 | -0.01 | – | 0.00 | 0.141 |
| Ferritin on Discharge | 18 | 416 | 18 | 779 | -249 | -611 | – | -19 | 0.029 |
| D Dimer on discharge | 14 | 0.5 | 17 | 0.8 | -0.33 | -1.06 | – | 0.08 | 0.115 |
| Procalcitonin on discharge | 17 | 0.05 | 18 | 0.05 | 0 | -0.03 | – | 0.03 | 0.980 |

The CP antibody level was available for 13 participants (mean 63.8 AU/mL± sd 46.8, median 54.5). The patients who required a ventilator and received CP had a mean antibody level of 84.95 AU/mL (SD 13.72, SE 7.9, N=3), whilst those that did not require mechanical ventilation had a mean of 57.48 AU/mL (SD 51.8, SE 16.4, N=10; p=0.24).

30% of patients who received early CP (less than 3 days from admission) required ventilation. None of the patients who received CP after 3 days from admission required ventilation. Patients who received early CP had a mean antibody level of 82 AU/mL (SD 23, SE 9.5, N=6). Those who received CP after 3 days had a mean titre 49 AU/mL (SD 58, SE 22, N=7) (p=0.06, rank sum test).

Two patients died in the control and one patient in the CP arm and their secondary outcome data were not included in Table 3

### Adverse Events

Three patients treated with plasma reported adverse events during the study that were not considered to be related to therapy: one with diarrhoea and vomiting that settled spontaneously; one with constipation and one desaturated transiently after the infusion.

## Discussion

In this pilot study, the CP group appeared to have a higher systemic inflammatory response shown by the increased ferritin levels and more patients with chronic lung disease were included, a known risk factor for more severe disease ^12^; however, respiratory indices did not differ between the groups. While the CP therapy group had fewer patients deteriorating to ventilation (primary outcome), and the duration of ventilation was less, this did not differ significantly from the standard therapy group. These results are in accordance with a larger study done in China by Ling Li et al, that was stopped prematurely due to the decrease in COVID 19 cases ^7^; however, as shown in this pilot, it is likely that trial was underpowered as an estimated sample size of over 822 in each group may be required to show a difference.

Steroids have been shown to be effective in COVID-19 ^13^; however, at the time of the study this evidence was not known and hence steroids were used at the discretion of the physicians. Steroids were used on 3 patients on standard therapy, none of the CP patients received steroids, but the sample size was inadequate to justify a hypothesis test. Of note, in the study by Ling Li et al, 45.6% CP arm and 32.7% of control arm were given steroids that may have masked the CP effect^7^, and in addition, the impact of Chinese herbal medicines (used in over 50% of patients) is unclear in COVID-19 ^7^. In this study, no subject received Remdesivir, but Hydroxychloroquine, Ribavirin and Lopinavir/ritonavir were used, though to date they have not been reported to be effective in COVID-19 infection ^14 15 16^. This pilot in accord with others ^7 17 18^ indicated that CP therapy was safe with only three transient adverse reactions being recorded. The safety of plasma was assessed in an observational study in 20,000 hospitalized patients in the US and it reported low incidence (<1%) of serious adverse events^18^. Sanfilippo et al highlighted that plasma transfusion can be harmful in COVID19 as plasma contains procoagulant factors, and COVID19 represents a unique scenario as they tend to have an increased risk for thrombosis^19^. The prothrombotic risk with plasma transfusion was not investigated in our study nor in other studies, and it has been suggested recently that the potential harm of the non-immune components of convalescent plasma should be studied, especially the prothrombotic risk.^20^

Patients who received early CP (less than 3 days from admission) had a higher mean antibody level in the transfused plasma, than those that received CP after 3 days, though the difference was not significant. A previous study reported that patients who received early CP and with higher antibody titres showed a better clinical outcome^8^; however, this pilot was not adequately powered to test the effect of early plasma in comparison to late plasma administration.

The CP collected from donors showed variations in mean antibody level, some extending to very low levels. The variability in the antibody level can have an effect on the effectiveness of plasma and can underestimate the effect of CP in this trial. The FDA have given recommendations to use CP with high antibody titre only ^21^, to prevent transfusion of CP with low antibody levels that can be ineffective.

Our trial result was also in agreement with a recent randomized trial conducted in India in 464 adults with COVID19. Agarwal et al mentioned that convalescent plasma was not associated with a reduction in progression to severe covid-19 or all-cause mortality. A subgroup analysis showed no difference in the outcome even after stratifying on the presence neutralizing antibody levels (> 1:20). ^22^

A randomized trial comparing convalescent plasma with standard of care therapy in patients hospitalized for COVID-19 in the Netherlands was stopped early after observing that more than 79% of patients randomized to the plasma arm had median antibody titres comparable to the plasma donors, prior to receiving the plasma transfusion. There was no significant difference in the median neutralizing antibody titre between recipients and donors, despite patients being randomized within 10 days of symptom onset. This may indicate that measuring the antibody titre of patients is important in order to select those patient with low a antibody titre that might benefit from CP transfusion. ^23^

The main limitations of this study were that it was pilot whose main purpose was to guide the design of a definitive study of CP therapy, and as a consequence it lacks the statistical power to conduct outcome hypothesis tests. Determination of the optimal antibody titre from the donors should also be undertaken that was not done in this study, as well a measurement of antibody titre in the recipients before and after the infusions. Moreover, the quantification method for the antibody levels could have been improved and using the neutralizing antibody titre would have been more appropriate; however, at the time of the study, an authorized neutralizing antibody titre test was not available. Furthermore, the antibody titres were also not measured for our patients on randomization.

This pilot study showed that it is feasible to perform a phase 3 randomized controlled trial on a larger sample size to determine the effect of plasma, using the current protocol. For a definitive study, the following are suggested modifications 1) Including only plasma with high titre antibody, (defined as ≥250)^21^; 2)Include only recipients with low antibody titre levels; 3)Serial monitoring of PaO2:FiO2 Ratio in patients; 4)Using of “hard” composite outcome of death or severe ARDS denoted by PaO2:FiO2 Ratio <100; 5)Studying safety endpoint regarding thrombosis in patients using plasma.

In conclusion, there were no significant differences in the primary or secondary outcome measures between CP and standard therapy though fewer patients required ventilation and for a shorter period of time. The study showed that CP therapy appears to be safe and it is feasible to perform a definitive phase 3 clinical trial using this study protocol.

## Supporting information

Appendix: Study protocol

## Data Availability

Data will be made available upon reasonable request

## Acknowledgements

We are very grateful for the support in this trial by our fellow colleagues from physicians and nurses who have worked hard throughout the pandemic and specially during the trial period.

Dr Omar Yaghi, Dr. Bandar Alnajdi, Dr. Hajer Alahmadi, Dr. Reem Almehzae, Dr. Layla Al Mutawa, Dr. Mahmood Tooq, Nurse Shafeeqa Hasan Yousif, Nurse Abdulla Mohammed Al-Ghadban, Nurse Hasan Hameed Hasan, Nurse Hamad Yusuf Habib, Nurse Mohamed Habib Hussain, Nurse Ali Abdulhadi Ali, Dr. Abdulla Alfadhel, Dr. Dana Alsharqi, Dr. Fatema Nabeel Nedham, Dr. Rashid Al Hasan, Dr. Ahmed Al Ansari, Dr. Maha Bukamal, Dr. Manar Al Sulaiti, Dr. Alya AlDoseri, Nurse Ghaida Fareed Ahmed Hasan, Nurse Elham Alfaraj, Nurse Abdulla Aman, Nursee Mariam Burshaeed, Nurse Manar Aldoseri, Nurse Ammar AlAthamneh, Nurse AlAnood Abdulrahman, Nurse Mariam Hasan Mohammed, Nurse Fatima Basheer, Nurse Ebtisam Meshal Saleh, Nurse Tareq Hasan Abdulrahman, Nurse Mariam Saleh Barshed Nurse Shouq Nader Hamada

We would also like to extend our gratitude to all health care workers and first responders worldwide and especially in Bahrain, who work day and night to protect the people and fight the virus.

