## Appendix: Study protocol for "Randomized controlled trial of convalescent plasma therapy against standard therapy in patients with severe COVID-19 disease"

الحملة الوطنية لمكافحة فيروس كورونا (COVID-19)

RCSI

RCSI DEVELOPING HEALTHCARE LEADERS WHO MAKE A DIFFERENCE WORLDWIDE

***Full Title: Convalescent plasma therapy for COVID-19 patients: a prospective randomized trial***

**Short title:** Plasma trial in COVID -19 patients


**Principal Investigator: Dr Manaf Al Qahtani**

**Co-Investigators:** Dr Manal Abduljalil Al Sayed, 36944559

Dr Abdulrahman Almadani 36995665

Dr Abdulkarim Abdulrahman 33008525

Dr Salman Yousif Alali 39788500

Dr Amal Hejab 33411064


Managing consultant of patients and randomizing physicians will be added as an amendment to ethics as the need arises

**Medical Monitor:** Mr. Aslam

**Coordinating Centers:**

- Hereditary blood Disorder Centre – Salmaniya Medical Complex
  Address - Rd No 2904, Manama. PI, Dr Alaa Al Zamrooni

- Bahrain Defense Force Hospital Blood Bank
  Address – Wali Al Ahd Hwy – Riffa. PI, Dr Manal Alsayed

- Bahrain Defence Force Royal Medical Services, Military Hospital
  Address – Wali Al Ahd Hwy – Riffa. PI, Dr Abdulrahman AlMadani

- Royal College of Surgeons in Ireland – Bahrain
  Address - Building No. 2441, Road 2835, Busaiteen 228. PI, Professor Stephen Atkin

### 1. Introduction/Background

Coronavirus disease 2019 (COVID-19) is caused by the severe acute respiratory syndrome coronavirus 2 (SARS-CoV-2) and has developed into a pandemic with serious global public health and economic sequelae. As of March 30, 2020, over 750,000 cases have been confirmed worldwide leading to over 34,000 deaths (https://coronavirus.jhu.edu/map.html). There is no current vaccine available, but there have been a number of reports of medication such as hydroxychloroquine having antiviral properties with efficacy against SARS-CoV-2.

Plasma therapy using convalescent plasma has been shown to be effective in severe acute respiratory syndrome (1), Ebola virus infection (2) and in H1N1 influenza (3). More recently there has been a report of the use of convalescent plasma in the treatment of 5 ventilated COVID-19 patients with the suggestion of expedited recovery as the patients improved 1 week after the transfusion (4). However, this was not a clinical trial and the patients were on other antiviral medication.; therefore, there is a need to undertake such a trial to see if deploying plasma with SARS-CoV-2 neutralizing antibody has utility in managing patients infected with COVID-19 in respiratory distress.

### 2.Objectives

#### Primary objective

The objective of this pilot study is to compare plasma therapy using convalescent plasma with antibody against SARS-CoV-2 to usual supportive therapy in COVID-19 patients with pneumonia and hypoxia, and to determine if the clinical course is improved. The difference between groups will allow an effect size to be determined for a definitive clinical trial

#### Research Question

Could using convalescent plasma transfusion, from recovered COVID19 patients with antibody against COVID-19 be beneficial in treatment of COVID19 patients with hypoxia and pneumonia, in order to avoid or delay the need for ventilation?

### 3.Study Design

This is a prospective, interventional and randomized open label trial involving 40 patients with COVID-19 who are in respiratory distress, with the criteria that all require oxygen therapy and have radiological evidence of pneumonia, 20 of whom will receive a single transfusion of convalescent patient plasma plus routine care, compared to 20 COVID-19 patients who will receive routine care alone.

**Study Arms:**
1. Control group:  local standard of care which include antivirals and supportive care
2. Intervention group: convalescent patient plasma plus routine local standard of care

**Outcomes**
Primary outcome is 1) Requirement for ventilation (invasive or noninvasive)
Secondary outcomes: 1) Improvement in biomarkers 2) Mortality rate 3)Time to viral clearance (using Ct values)

**Identification of suitable plasma donors among patients recovered from COVID-19**.
Patients who have recovered from COVID-19 and have been discharged from hospital will be approached to see if they will volunteer to be donors

**Criteria for donors**
1. Able to give informed consent
2. Male or nulliparous women(women will need a pregnancy test except for postmenopausal women)
3. At least 14 days after discharge
4. Clinically asymptomatic
5. PCR COVID-19 negative from respiratory tract
6. Patients above the ages of 21 (with no upper age)
7. Body weight more than 50kg
8. At least one week since last glucocorticoid usage.
9. Meet all donor selection criteria employed for routine plasma collection and plasmapheresis procedures at the collection center

Following informed consent, the potential donor will be assessed for suitability to donate blood or plasma through the clinically established donor selection process that includes general health criteria such as such as weight, medical and social (i.e. behavioral risk factors) history, basic physical examination and hemoglobin estimation. One unit of blood will then be taken for transfusion. Donor confidentiality will be maintained throughout and potential donors will be informed that there will be no payment to them for their plasma donation. A minimum period of 12 weeks will occur in the event of a future blood donation.

Pre-donation testing should include:

1. ABO and RhD grouping,
2. Blood screening tests for HIV, HBV, HCV, syphilis and other locally transmitted infections, as applicable.
3. Hemoglobin estimation.
4. Nucleic acid testing for SARS-CoV-2.
5. Titration of total COVID-19 antibodies and COVID-19 neutralizing antibodies (If not available, the presence of antibodies qualitatively will be assessed)

6. 3 ml plasma will be retained for viral neutralization experiments.

Storage of plasma units, inventory management and transportation

- Donated Blood unit will be tested according to the local blood bank protocol.
- Donated Convalescent Plasma will be stored as ‘Plasma Frozen Within 24 hours’ and stored for up to 12 months at or below -18$^0$ C in a separate controlled plasma freezer dedicated to the trial. Longer storage period might be considered for research purposes only.
- Both whole blood units and plasma units will be tagged as COVID-19 Donor using labels, separate codes and serial numbers to make them identifiable from others. The plasma container will be labelled as Caution: New Drug Limited by law to investigational use”.
- Sample from convalescent plasma will be stored for determining antibody titer at a later date once validated SARS-CoV-2 neutralizing antibody titer test is available.
- Plasma unit will be transported to COVID center in a special labelled refrigerator and will be arranged between hospitals blood banks.

**Selection of COVID-19 patients**
1. Able to give informed consent
2. COVID-19 patients who have tested positive for SARS-CoV-2, and who are hypoxic and requiring oxygen therapy (Oxygen saturation of less than or equal 92% or PO2 < 60mmHg on ABG)
3. Radiological evidence of pneumonia (any infiltration on CXR or CT scan.)

**Contraindications:**
1. History of allergy to plasma, sodium citrate or methylene blue.
2. History of autoimmune disease or selective IGA deficiency.

**Side effects**
The major potential side effect will be that of a transfusion reaction that may be manifest with increased fever, hypotension and skin rash. This will be minimized by the ABO and RhD grouping that will be taken for each patient.

**Convalescent plasma administration**
ABO and RhD matched plasma units will be selected for transfusion and transfused to the COVID-19 patients using standard clinical transfusion procedures.
The dosage of convalescent plasma will be 400 mL, given as 200ml over 2hrs in 2 consecutive days.

**Study Schedule**

**1: Patient characteristics**
- Patient confirmed to be SARS-CoV-2positive by PCR
- Patients able to give informed consent
- Patients who are hypoxic and requiring oxygen therapy

- Consent, inclusion and exclusion criteria. Patients will be assessed for the inclusion and exclusion criteria of the study by the consultant and consented by a doctor on the clinical team.
- Patient identifiers, admission date, age and sex
- Patient characteristics (yes/no): current smoking, diabetes, heart disease, chronic lung disease, chronic liver disease, chronic kidney disease, asthma, HIV, tuberculosis
- COVID-19 oxygen therapy being given, oxygen saturation on room air, oxygen flow.
- Anthropometric measurement –BMI, blood pressure, temperature, respiratory rate
- ECG
- Baseline bloods: Biochemical profile (BCP), full blood count (FBC), CRP, PCT, LDH, Troponin, Ferritin, D-Dimer, BNP, lactate
- Baseline ABG on room air, on enrolment
- Baseline PaO2 value
- Chest Xray prior to starting plasma transfusion
- ABO and RhD blood grouping and cross-matching
- Baseline SARS-CoV-2 Swab PCR with CT Value (indirect assessment of viral load) through Nasopharyngeal swab.
- Randomization into one of the 2 groups

**2: Daily recording in case report form for 10 days or until discharge**

- blood pressure, temperature, respiratory rate
- Oxygen flow rate and oxygen device for oxygen saturation
- Full blood count(Hb, WBC, Neutrophil count, Neutrophil percent (%),Lymphocyte count, Lymphocyte percent (%),Platelets) , Biochemical profile(Na, K, Urea, Creatinine, LDH), Liver function tests(ALT,AST,ALP,GGT), CRP, PCT, LDH, Troponin, Ferritin, D-Dimer, BNP, lactate
- Date of plasma infusion
- Amount of plasma infusion
- Adverse Event or Adverse Reaction
- Patient’s current medications
- Date of Intubation
- Ventilator Setting if on ventilator

**3: Intermittent recording in case report form for 10 day or until discharge**

- Chest Xray 1 day after completion of plasma therapy or standard treatment and on Discharge
- ABG and PaO2 values after completion of plasma therapy or standard treatment on day 1,2,3 and then every 3 days and on discharge
- SARS-CoV-2 Swab PCR with CT Value , day 1 and 3 after plasma transfusion or standard treatment, every 7 days, and on discharge

*Criteria for discontinuation of the study*.
The study will be discontinued based on any adverse reactions such as transfusion reactions or may be withdrawn by the Ministry of Health decision on safety, or if any superior drug/intervention has been identified and is available

### 4.Materials and methods:

#### Subject selection

40 sequential patients, both males and females, diagnosed with COVID -19 and admitted to hospital as an inpatient, requiring oxygen therapy with radiological evidence of pneumonia will be recruited, 21 years of age or older, with no upper age limit (particularly as the elderly are more at risk of an increased mortality). Whilst the Chinese data suggested that males were at higher risk of increased severity than females, the primary aim of the study is to show efficacy and therefore stratification by gender will not be undertaken.

All subjects SARS-CoV-2 positive on oxygen therapy and radiological evidence of pneumonia will be offered the opportunity of entering into the clinical trial

*Patient population*

Number of subjects planned to be randomised: 40

Number of subjects expected to complete the trial: 40

SARS-CoV-2 positive patients on oxygen therapy and radiological evidence of pneumonia will have the study explained and informed consent will be undertaken at the bedside. Whilst this study is particularly addressing those patients who are English or Arabic speakers, this will not preclude those who have limited language ability in English/Arabic and a translation service will be provided. Patients will then be assessed for exclusion criteria of the study and following randomization then treatment will be instituted

#### Inclusion Criteria

1. COVID-19 diagnosis
2. Hypoxia, (Oxygen saturation of less than or equal 92% or PO2 < 60mmHg on ABG) and patient requiring oxygen therapy
3. Evidence of infiltrates on Chest Xray or CT scan

2. Able to give informed consent
3. Patients between the ages of 21 and above with no upper age.

#### Exclusion criteria

1. Patients with mild disease not requiring oxygen therapy
2. Patients with normal CXR & CT scan
3. Patients requiring ventilatory support
4. Patients with a history of allergy to plasma, sodium citrate or methylene blue, or those with a history of autoimmune disease or selective IGA deficiency.

### 5. Randomisation

The study will randomise the patients to one of 2 groups randomised by computer. The number of patients in each group will be the same and they will have an equal chance of being in either arm of the study. Randomisation code will be provided by RCSI Bahrain to be administered in the local environment.

### 6.Withdrawal of Subjects

Once the plasma transfusion is given then it will not be possible to withdraw medication, and if the patient chooses to withdraw from the study their clinical data will still be included in the intention to treat analysis undertaken on study evaluation.

### 7.Data and Specimen Management and Quality control and quality assurance

Patients will be assessed clinically, and their vital signs and progress will be documented in the clinical notes and in the clinical report form

Should there be any missing data then missing values will be reported in the baseline table but won't be considered in the analysis.

### 8.Samples storage:

Serum samples will be sent to the laboratory processing and storage at -80C for future cytokine and antibody analysis.

Study I.D code will be assigned for each subject on day of recruitment. Samples will be stored and linked to the study I.D code. Data will be entered by the assigned CRC or any assigned study staff and then stored using a secured data base server. Each specimen will have a code only on the tube. No name, subject I.D, date or other identifying information will be on the specimen.

The Lead PI and the researchers assigned by him will have access to the stored data/specimens. Only the Lead PI and his delegated authority working on this study will be eligible to obtain the data/specimens from the participants.

Under no circumstances will any data or specimens be uncoded. After each study visit the collected data will be entered on data entry program on secure computer, the password to this computer will only be in the custody of the assigned CRC. The location of this computer will be at the BDF. The hard copy of the data will be kept in a locked cabinet, the assigned LPI or his delegated authority, will be the only personal having the keys. The collected specimens from the subject each session will be coded immediately then put in the laboratory freezer for future batch analysis.

### 9.Sample Size

There are currently no robust studies in man that would allow for a definitive power calculation and therefore this study can be considered a pilot study with the outcome of providing an effect size that would allow the powering of a definitive study. Power and sample size for pilot studies has been reviewed by Birkett and Day(9). They concluded that a minimum of 20 degrees-of-freedom was required to estimate effect size and variability and hence 20 patients per group have been included in this study; dropouts are unlikely to be an issue in this study.

### 10.Statistical Analysis

Baseline continuously distributed data will be presented as means and standard deviations unless the data is non-normal in which case the data median (25th/75th centiles) will be determined; categorical data by n (%). Given randomisation to treatment, baselines will not compared statistically (10-12). Intention to treat analysis will be undertaken. Within-group differences will be shown for each treatment group separately by a mean and a standard deviation (SD). Between-group comparisons will be performed using Student's t testing if the data is normally distributed, and non-parametric testing if it is not. For all statistical analyses, a two-tailed P <0.05 will be considered to indicate statistical significance. Time to event analysis and logistic regression will be used to analyse outcomes. Statistical analysis will be performed using the STATA statistical computer package (StataCorp. 2013. Stata Statistical Software: Release 13. College Station, TX: StataCorp LP, USA).

### 11.Provisions to Monitor the Data to Ensure the Safety of Subjects

Safety considerations/Patient safety:

Safety monitoring will be undertaken as a part of the clinical trial. A data safety monitoring board will be convened to review safety data on a weekly basis during the conduct of the study.

The major potential side effect will be that of a transfusion reaction that may be manifest with increased fever, hypotension and skin rash. This will be minimized by the ABO and RhD grouping that will be taken for each patient.

An interim review and report to the clinical trial committee will be undertaken when 20 subjects have been recruited

**Definitions**

| Term | Definition |
|---|---|
| **Adverse Event (AE)** | Any untoward medical occurrence in a participant to whom a medicinal product has been administered, including occurrences which are not necessarily caused by or related to that product. |

| | |
|---|---|
| **Adverse Reaction (AR)** | An untoward and unintended response in a participant to an investigational medicinal product which is related to any dose administered to that participant.<br><br>The phrase "response to an investigational medicinal product" means that a causal relationship between a trial medication and an AE is at least a reasonable possibility, i.e. the relationship cannot be ruled out.<br><br>All cases judged by either the reporting medically qualified professional or the Sponsor as having a reasonable suspected causal relationship to the trial medication qualify as adverse reactions. |
| **Serious Adverse Event (SAE)** | A serious adverse event is any untoward medical occurrence that:<br><br>• results in death<br>• is life-threatening<br>• requires inpatient hospitalisation or prolongation of existing hospitalisation<br>• results in persistent or significant disability/incapacity<br>• consists of a congenital anomaly or birth defect<br>Other 'important medical events' may also be considered serious if they jeopardise the participant or require an intervention to prevent one of the above consequences.<br><br>NOTE: The term "life-threatening" in the definition of "serious" refers to an event in which the participant was at risk of death at the time of the event; it does not refer to an event which hypothetically might have caused death if it were more severe. |
| **Serious Adverse Reaction (SAR)** | An adverse event that is both serious and, in the opinion of the reporting Investigator, believed with reasonable probability to be due to one of the trial treatments, based on the information provided. |
| **Suspected Unexpected Serious Adverse Reaction (SUSAR)** | A serious adverse reaction, the nature and severity of which is not consistent with the information about the medicinal product in question set out:<br><br>• in the case of a product with a marketing authorisation, in the summary of product characteristics (SmPC) for that product<br>• in the case of any other investigational medicinal product, in the investigator's brochure (IB) relating to the trial in question |

**Reporting adverse events**

Adverse events will be reported in accordance with the local procedures to the REC of NHRA

The following information will be reported: Study name; patient identification, including subject number, initials, sex, age; event (preferably diagnosis); trial drug Reporter Causality Outcome

**Determination of causality of an adverse reaction**

Probable - Good reason and sufficient documentation to assume a causal relationship.

Possible - A causal relationship is conceivable and cannot be dismissed.

Unlikely - The event is most likely related to aetiology other than the trial product.

**Adverse Event reporting period**

The AE reporting period for this trial begins as soon as patients have consented to the trial and ends 30 days after the patient's final study medication visit.

The health status of subjects will be checked at each study visit. The investigator will record all directly observed AEs and all AEs spontaneously reported by the trial subject.

A pre-existing condition is a disorder present prior to the patient entering the trial and does not need to be reported as an AE unless the condition worsens, or episodes increase in frequency during the AE-reporting period. A pre-existing condition will be documented at the screening or baseline study visit.

All adverse events (serious and non-serious) will be recorded by the investigator in patients data collection forms (CRFs) using R&D's adverse event report form. All adverse events will be recorded by the investigator in patients' medical records/notes.

All AEs will be followed-up by investigators until the event has resolved or a decision has been taken for no further follow-up.

If a clinically significant abnormal laboratory value occurs, this abnormality will be recorded as an adverse event/reaction.

**Reporting serious adverse events**

Investigators will notify the sponsor of serious adverse events within 24hrs of becoming aware of the event, together with the REC.

The sponsor will report fatal or life-threatening SUSARs within 5 days and follow-up information within a further 8 days. The investigator will report fatal or life-threatening SUSARs to the IRB within 5 days and follow-up information within a further 8 days by following the request on the Serious Event Initial and Follow-up report forms. The investigator will send all other SUSAR reports to the IRB within a maximum of 15 days.

**End of Trial:**

The end of the trial will be the date of the last study point of the last patient undergoing the trial.

An end of trial declaration form will be submitted to RCSI Bahrain/ BHF and NHRA REC within 90 days from completion of the trial and within 15 days if the trial is discontinued prematurely. A summary of the trial final report/publication will be submitted to the LREC within 1 year of the end of trial.

Serious adverse events will be reported to the sponsor within 24 hours. All other adverse events will be reported within 5 days.

**Potential Benefits to Subjects**

Being a part of the study will not guarantee benefits to subject or others. However, possible benefits might be that the subject's COVID-19 respiratory distress will improve and mitigate the infection within a 1 week period that may lead to the early discontinuation of oxygen administration and prevent the need for ventilatory support. Overall the research may benefit future patients with COVID-19 with severe respiratory distress.

**Indemnity/clinical trial insurance**

Negligent indemnity will be provided for by the ministry of health and any adverse effects on the patient medical status will be managed by the ministry.

**Confidentiality**

The study will abide to European GDPR legislation for maintaining confidentiality of data

**Ethical permissions**

The study will be performed subject to a Local REC favourable opinions from RCSI Bahrain and Salmanyia hospital, and from the National REC NHRA.

**Research Governance**

This study will be conducted in line with the International Conference for Harmonisation of Good Clinical Practice (ICH GCP) guidelines; the Research Governance Framework for Health and Social Care.

#### Data handling and record keeping

Dr Manaf will act as the data custodian and is responsible for the storage, handling and quality of the study data.

Data will be collected in the case report form to allow for cross referencing to check validity.

Study documents (paper and electronic) will be retained in a secure (kept locked when not in use) location during and after the trial has finished. All essential documents including source documents will be retained for a period of 5 years after study completion (last patient, last study point). A label stating the date after which the documents can be destroyed will be placed on the inside front cover of the case notes of trial participants.

#### Access to Source Data

Monitoring, audits, and REC review will be permitted and provide direct access to source data and documents. The Lead PI and the researchers assigned by him will have access to the stored data/specimens. Only the Lead PI and the researchers assigned working on this study will be eligible to obtain the data/specimens from the participants during data collection.

### 12.Dissemination of results and publication policy

The main findings will be widely reported urgently in the literature as appropriate. It is anticipated that this study will be reported at national and international conferences along with a number of papers in peer reviewed academic journals. The study will be registered at clinicaltrials.gov. Reporting of the study findings will be in accordance with the CONSORT statement (Consolidated Standards of Reporting Trials http://www.consort-statement.org/).

### 13.References
